# A prospective observational study on BBV152 coronavirus vaccine use in adolescents and comparison with adults- first real-world safety analysis

**DOI:** 10.1101/2022.04.08.22273634

**Authors:** Upinder Kaur, KL Anju, Mayank Chauhan, Aditi Joshi, Sangeeta Kansal, Vaibhav Jaisawal, Kishor Patwardhan, Sankha Shubhra Chakrabarti

## Abstract

**Background:** The BBV152 COVID-19 vaccine (COVAXIN) has recently been approved for adolescents. We provide the first real world safety data of COVAXIN use in adolescents and compare this with adults.

**Methods:** A prospective observational study is being conducted since January 2022. Enrolled adolescents and adults were contacted telephonically after 14 days of receiving the BBV152 vaccine. Primary outcome was vaccine safety assessed as rates of adverse events following immunization (AEFI). Severity grading of AEFIs was done using the FDA scale.

**Findings:** A total of 698 adolescents and 326 adults were enrolled. AEFIs occurred in 36.3% adolescents after first and in 37.9% after second dose. Systemic involvement was seen in 15-17% adolescents. Injection site pain and fever were the common AEFIs. Majority of AEFIs were mild-moderate. Severe and atypical AEFIs were observed in 0.9% and 0.6% adolescents respectively. Majority of AEFIs recovered in 1-2 days. In >2% adolescents, AEFIs were persisting at 14-day follow-up since the second dose. No difference in AEFI incidence and patterns was observed between adolescents and adults. Regression analysis showed females and those with history of allergy to be respectively at 1.5-times and 3-times increased risk of AEFIs among adolescents.

**Interpretation:** COVAXIN carries an overall favorable short term safety profile in adolescents. The observed AEFI rates in adolescents are much lower than that reported with mRNA vaccines. Female adolescents and those with history of allergy need watchfulness. With some AEFIs persisting at 14 days, a longer follow-up is recommended to strengthen the safety data of these vaccines.

**Funding:** No funding support

## 1. Introduction

Vaccines based on novel and pre-existing platforms have been licensed for use to curtail the Coronavirus disease-2019 (COVID-19) pandemic. These include messenger RNA (mRNA) based vaccines, viral vectored vaccines, and inactivated severe acute respiratory syndrome coronavirus-2 (SARS-CoV-2) vaccines. Initially high-risk groups were prioritized for vaccination, and later on the programme was extended to the general population. The World Health Organization (WHO) recommends extension of vaccination to children 5-17 years of age once the high-risk population has been adequately vaccinated.^1^ Pfizer’s mRNA based BNT-162b2 vaccine was the first to get Food and Drug Administration (FDA) and European Medicines Agency (EMA) approval for vaccinating children against COVID-19.^2^ The inactivated CoronaVac in China has also been granted emergency use approval for children and adolescents.^3^ In India, COVISHIELD based on the ChAdOx1-nCoV-19 chimpanzee adenoviral platform and COVAXIN (BBV152), an inactivated SARS-CoV-2 vaccine were mostly used for mass vaccination of the adult population. Of these, COVAXIN has recently been granted emergency use authorization for use in adolescents 15-18 years of age. There is paucity of data on safety profile of inactivated vaccines specific to the adolescent subset. The only evidence available till date is a pre-print of an open label phase 2/3 trial of the BBV152 vaccine by the manufacturer Bharat Biotech which enrolled only 175 adolescents. The vaccine was shown to have a favorable short term safety profile.^4^ Since the performance and safety of vaccines might differ in larger population samples in the real world and also on long term follow-up, we decided to conduct a prospective observational study of the BBV152 vaccine (COVAXIN) in adolescents aged 15-18 years. Here we provide the first interim real world safety data of COVAXIN in adolescents. A comparative analysis of short-term vaccine safety in adolescents and adults is also presented.

## 2. Materials and Methods

### 2.1. Study design and setting

This is a prospective observational study conducted in a tertiary hospital of north India. The study started in January 2022 and with a target of one year follow-up of all participants, is expected to be continued till May 2023. Adolescents visiting the vaccination center at the time of either first or second dose were recruited in the study. By the time the study started, a subset of adolescents visiting the centre had already received their first dose, and they were recruited during their second dose. Information regarding AEFIs during first dose received 4 weeks back was obtained from these participants and their accompanying guardians at the time of second dose and was recorded in the pre-designed case report form. Adult participants in the study were recruited while visiting the vaccination center at the time of either their first or second or booster dose. As per the Ministry of Health and Family Welfare (MoHFW) guidelines, adults were eligible for booster if they had received the second dose 9 months back.^5^ Some of the adults receiving the second dose had received the first dose more than 3 months back. Because of the significant time lag and the possibility of recall bias, retrospective evaluation of AEFIs was not performed in adults who were enrolled at the time of either second or booster dose. Adult participants were only monitored prospectively to assess the AEFIs.

Here we report the first short term safety results of all participants-adolescents or adults who successfully completed at least fourteen days of follow-up post any dose of the BBV152 vaccine (COVAXIN). The authors UK, SSC and VJ had access to the complete data.

### 2.2. Study participants

All individuals who received COVAXIN in the study centre during the period of enrolment were included in the study. Participants were adolescents of 15-18 years age and adults ≥ 19 years of age. Informed consent was taken from the adults. In case of adolescents, informed consent was taken from the accompanying guardian along with written assent of the adolescent. Adolescents who visited the centre without guardians were excluded as were the vaccinees who refused to provide consent/assent.

### 2.3. Safety analysis

Adverse events following immunization (AEFIs) were recorded at 14-day post vaccination through telephonic interview and the following detailed data for safety analysis were extracted.

∘ Incidence of AEFIs
∘ Type and pattern of AEFIs (Medical dictionary for regulatory activities, MedDRA low level terms and system organ class terminology used)
∘ Distribution of AEFIs with respect to age and sex
∘ Outcomes of AEFIs
∘ Interventions done to manage AEFIs
∘ Seriousness of AEFIs as per Food and Drug Administration (FDA) definition
∘ Severity of AEFIs for local adverse events (AEs), systemic AEs, and vital signs. These were recorded as per FDA severity grading scales
∘ AEFIs requiring hospitalization.
∘ Any vaccine-disease interaction resulting in AEFI
∘ Any vaccine-drug interaction resulting in AEFI

### 2.4. Vaccination procedure and enrolment in study

Participants receiving any dose of COVAXIN were recruited for the present study. COVAXIN is administered intramuscularly in the deltoid at the dose of 0.5 mL as a two-dose schedule, with an interval of 4-6 weeks between the first and second dose. Since mid-January 2022, the booster dose of vaccine was recommended for adults with a gap of 9 months from the second dose.^5^ Each dose of 0.5 mL contains 6µg of whole-virion inactivated SARS-CoV-2 antigen (Strain: NIV-2020-770), and the other inactive ingredients such as aluminium hydroxide gel (250 µg), TLR 7/8 agonist (imidazoquinolinone) 15 µg, 2-phenoxyethanol 2.5 mg, and phosphate buffer saline up to 0.5 mL. Post vaccination, all recipients are routinely monitored at the study site for 30 minutes. The enrolled participants were informed of the adverse events expected after vaccination and of the study procedure. They were contacted by phone at day 14 post vaccination. Specifically, they were questioned about local site symptoms such as pain, erythema, swelling, tenderness, and any limitation of physical activity. Also, enquiry was made about the occurrence of systemic events such as fever, fatigability, myalgia, arthralgia, headache, nausea, vomiting, diarrhoea, rash, chest tightness and dyspnoea. With the third wave of the COVID-19 pandemic ongoing, individuals were instructed for RT-PCR based nasal or oropharyngeal swab test for SARS-CoV-2 in any event of developing COVID-19 like symptoms.

### 2.5. Ethical permission

The study started after obtaining permission from the Ethics Committee of the Institute of Medical Sciences, Banaras Hindu University, and written informed consent was taken from the adults. In case of adolescents, informed consent was taken from the guardians along with the written assent of adolescents.

### 2.6. Data sources/measurement

Data related to demography and medical history was recorded in a pre-designed case report form. The medical history included history of SARS-CoV-2 positivity at any time in the past, existing co-morbidities, concurrent drug history and history of allergy to any known stimuli. For adolescents visiting the centre for second dose, information regarding AEFIs during the first dose was recorded in the case report form. For all participants, the data recorded included severity of AEFIs, interventions required for the management of AEFIs, outcomes of AEFIs and time to complete recovery.

### 2.7. Sample size

The available safety data from controlled settings shows the AEFI rate to vary from 14-21% in the general population. The primary outcome of the study was to evaluate the safety profile of COVAXIN in adolescents. Considering a 15% rate of occurrence of AEFI, margin of error of 4%, and 10% rate of drop out, the expected sample size for the present study was calculated to be 352. The study also aimed to compare the vaccine safety in adolescents with adult population. Considering the feasibility concerns, it was decided to enrol at least 1000 participants with adolescent:adult ratio of 2:1. The enrolment was stopped after 1024 participants were recruited.

### 2.8. Statistical analysis

For data such as incidence, severity, and outcomes of AEFIs, values were recorded as frequencies as well as percentages. Chi square test was applied for dichotomous variables such as sex, presence of co-morbidities and medications to find association between these potential risk factors and development of AEFIs. Comorbidities in adolescents being quite low in number, were analysed as a single composite variable (comorbidities) rather than separate entities like diabetes, hypertension, hypothyroidism etc. Variables with statistically significant association (P<0.05) on bivariate analysis or deemed to be clinically relevant were incorporated in the final regression model. In addition, a comparative AEFI analysis was done only between adolescents and adults enrolled while receiving the first dose. Others were excluded from this analysis to avoid recall bias. Results were analysed using SPSS version 16.

### 2.9. Role of funding source

This study has no funding support.

## 3. Results

**Figure 1** (as per STROBE guidelines) shows the recruitment of vaccinees for the present study. Of the 698 adolescents enrolled, 339 and 359 were respectively recruited at the time of first and second dose of COVAXIN. Of the 339 adolescents visiting the center for first dose, 28 were lost to follow up and among adolescents recruited at the time of second dose, 19 were lost to follow up. Thus, AEFIs at 14 days post-first dose were assessed in a total of 670 (i.e., 311+359) adolescents and AEFIs at 14 days post-second dose were assessed in 340 adolescents. A total of 326 adults were recruited in the study of whom 201, 113 and 12 were visiting the center for first, second and booster dose of COVAXIN respectively. Of the 326 adults enrolled, 31 were lost to follow up and AEFI data at 14-days post vaccination was available for the remaining 295. The baseline characteristics of the study participants are mentioned in **Table 1**.

**Table 1:** Baseline characteristics of enrolled adolescents and adults. DM: diabetes mellitus, PCOD: polycystic ovarian disease, SARS-CoV-2: severe acute respiratory syndrome coronavirus-2 [All percentages expressed with respect to total enrolled adolescents and adults respectively]

|  | **Adolescents**<br>**(N=698)** | **Adults**<br>**(N=326)** |  |
| --- | --- | --- | --- |
| **Female/Male** | 368/330 | 165/161 |  |
| **Age (Range in years)** | 15-18 | 19-82 (Median- 30) |  |
| **Body mass index (Mean ± SD)** | 19.7 ± 3.3 | 22.2 ± 3.7 | |
| **Co-morbidities** | 33 (4.7) | 66 (20.2) |  |
| Asthma | 8 | DM | 20 |
| Epilepsy | 4 | Hypertension | 20 |
| Skin disease | 7 | Hypothyroidism | 7 |
| Hypothyroidism | 4 | Migraine | 3 |
| PCOD | 2 | Asthma | 2 |
| Others | 8 | Kidney disease | 2 |
|  |  | Others | 12 |
| **History of allergy, N (%)** | 41 (5.9) | 27 (8.3) |  |
| **Prior SARS-CoV-2 infection, N (%)** | 39 (5.6) | 26 (7.9) |  |

**Figure 1.**
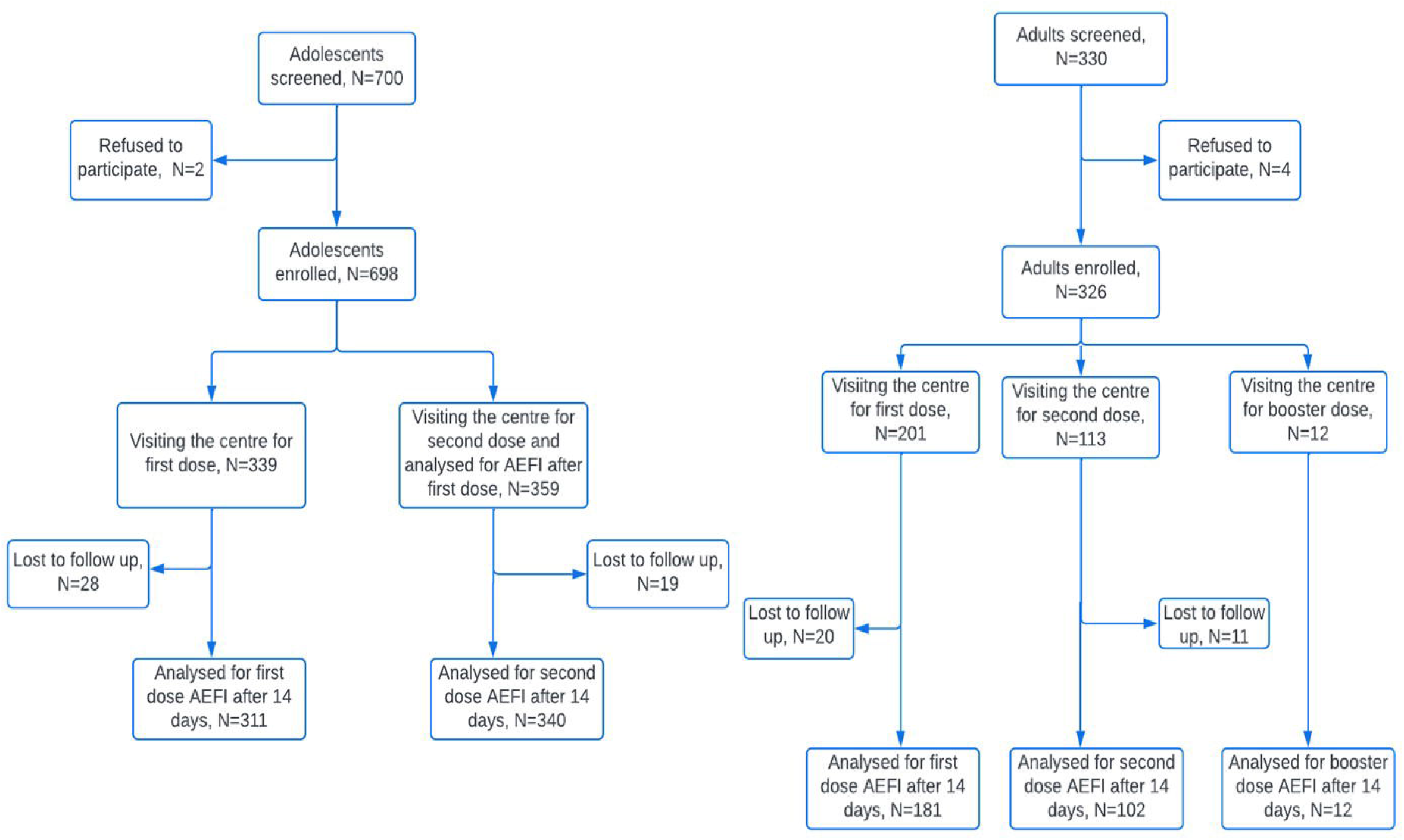
STROBE flowchart showing enrolment of adolescents and adults in the study.

### 3.1. AEFIs after first dose in adolescents

Among 670 adolescents for whom data was available, 244 reported systemic or local AEFIs (36.3%). Majority of AEFIs involved the injection site and were reported by 183 (27.3%) adolescents. Systemic involvement with or without local site involvement was reported by 102 (15.2%). Individually, injection site pain was the commonest AEFI reported (179, 26.7%) followed by fever (12.2%) and weakness (1.9%). Majority of AEFIs occurred within 24 hours of receiving the vaccine (n=233, 95.5% of all AEFIs). Most local site AEFIs were mild (152/183, 83.1%) and most systemic AEFIS were of ‘moderate’ grade (51/102, 50%). Altogether, six adolescents (0.9% of all adolescents) complained of systemic AEFIs of ‘severe’ grade and no ‘serious’ AEFIs were noticed. Till the date of telephonic follow-up, complete recovery from AEFIs was observed in all adolescents except three and time to recovery (TTR) varied from 0.5-7 days. Four atypical AEFIs were reported in four adolescents (0.6%). These included a case of severe menstrual bleeding with no endocrinal disturbance in past, a case of disturbing burning sensation in lower limbs in a male with insignificant past history, a case of recurrence of seizure within 5 days of first dose in a female with pre-vaccination uncontrolled seizure disorder and hypothyroidism within 3 weeks of first dose in a female with pre-vaccination neck swelling (**Table 2**). The latter two were reported to the study team at the time of second dose. The MedDRA SOCs of reported AEFIs are shown in **Figure 2**.

**Figure 2.**
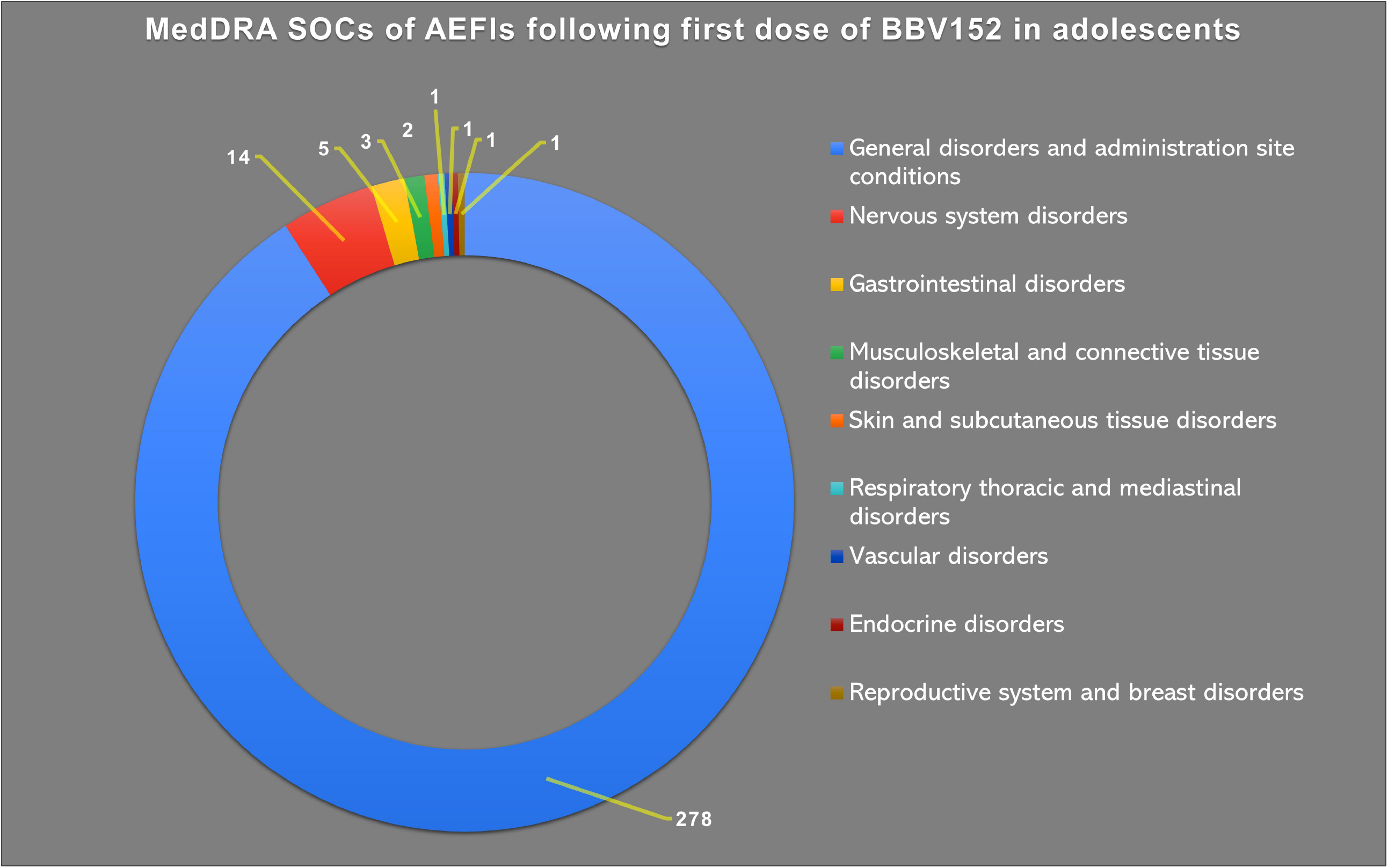
MedDRA SOCs of AEFIs following first dose of BBV152 in adolescents.

### 3.2. AEFIs after second dose in adolescents

Of 340 adolescents interviewed after the second dose, AEFIs were reported by 129 (37.9%). Among these, local site and systemic AEFIs were noticed in 96 (28.2%) and 60 (17.6%) adolescents respectively. Injection site pain was the commonest (28.2%) followed by fever (14.1%) and headache (3.2%). Majority of the local AEFIs were of ‘mild’ grade and majority of the systemic AEFIs were ‘moderate’. No ‘severe’ or ‘serious’ AEFIs were observed (**Table 2**). Majority of AEFIs occurred within 24 hours of the vaccine (94.6%) and recovery was seen in all except eight till the date of interview. TTR varied from 0.5-14 days. Three atypical AEFIs were reported in three adolescents (0.9%). These included one case each of epistaxis, recurrence of seizures in female with uncontrolled seizure disorder pre-vaccination, and aggravation of pre-vaccination skin allergy. The MedDRA SOCs of reported AEFIs are shown in **Figure 3**.

**Figure 3.**
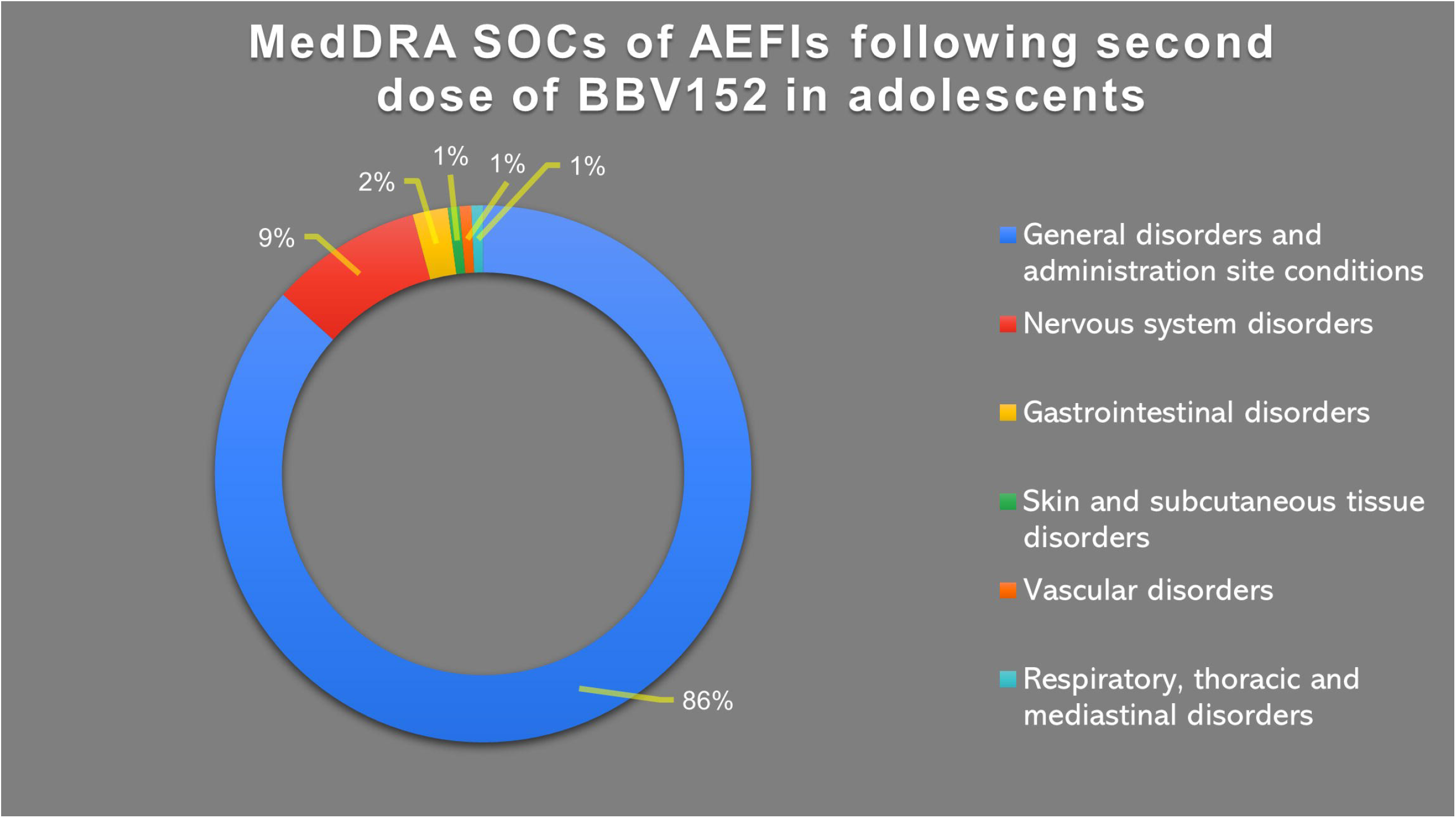
MedDRA SOCs of AEFIs following second dose of BBV152 in adolescents.

### 3.3. AEFIs in adults after first, second, and booster dose of COVAXIN

After excluding those lost to follow up, AEFIs after first dose were assessable in 181 adults and after second dose in 102 adults. AEFIs occurred in 93 (51.4%) after first dose and 38 (37.2%) after second dose of the vaccine (**Table 2**). Injection site pain and fever were common adverse events after both doses and majority of the AEFIs were mild-moderate. Only one severe and no serious AEFI was reported in adults. Among atypical AEFIs, epistaxis and heavy menstrual bleed were reported in one case each after the first dose. AEFIs after booster dose were reported in 41.7% and are mentioned in **Table 2**. AEFIs till the day of telephonic follow-up were persisting in 11 adults.

### 3.4. Comparative analysis between adolescents receiving first dose and adults receiving first dose

Direct comparison of AEFI occurrence was performed between adults and adolescents receiving the first dose (n=540). To ensure that the analysis is unadulterated by the dose of vaccine or recall bias, adults and adolescents only receiving the first dose and monitored prospectively were included. After excluding 48 participants lost to follow up, comparative analysis was conducted in the set of 492 participants. No statistically significant difference was observed in the local (P=0.75), systemic (P=0.40) or overall occurrence (P=0.90) of AEFIs between adolescents and adults.

### 3.5. Risk factors of AEFIs in adolescents

Bivariate analysis was performed to determine association between AEFIs occurring after first dose and potential risk factors. Variables selected were sex, history of previous COVID-19, presence of co-morbidities and history of allergy. A total of 502 adolescents were selected for analysis of risk factors following any dose, after excluding those who were lost to follow up. In unadjusted analysis, AEFIs occurred commonly in females and those with history of allergy. In adjusted analysis, however, only history of allergy was a significant determinant of AEFI occurrence. Adolescents with history of allergy were at 2.7 times higher risk of developing AEFIs compared to those with no history of allergy.

A separate regression analysis was performed to determine risk factors of AEFIs occurring after any dose, in adolescents who had completed their second dose. Those with no history of AEFI after the first dose but lost to follow up after second dose (n=15) were excluded. Thus, 344 adolescents were selected for this analysis. Variables selected for unadjusted analysis were sex, history of COVID-19, presence of co-morbidities and history of allergy.

In unadjusted analysis, statistically significant association was observed between the occurrence of AEFIs and female sex, as well as history of allergy. Association of both these factors remained statistically significant even on logistic regression. Females were at 1.6 times and those with history of allergy were at 3 times higher risk of development of AEFIs with respect to respective comparators.

## 4. Discussion

Post vaccination, serious adverse events such as the syndrome of thrombosis and thrombocytopenia, acute cardiac events, and new onset as well as flares of autoimmune diseases have been reported in adults.^6–8^ Myocarditis has been commonly observed with mRNA vaccines and was reported initially in adults. With recommendations to vaccinate adolescents, myocarditis, as assessed by passive surveillance methods, is being reported in this age group at an increased frequency.^9^ Rates of adverse events may be high in adolescents with underlying inflammatory diseases. In adolescents with juvenile rheumatic diseases, complications such as renal failure, pulmonary hemorrhage and flares of lupus have been reported with the BNT162b2 vaccine, each at the rate of 1 per 91 cases.^10^

The passive AEFI surveillance methods often suffer from underreporting. The actual risk of rare but severe adverse events needs evaluation from a prospective cohort-based design. To fill this gap, the present study was planned to generate evidence on short-term and long-term safety profile of COVAXIN in adolescents who are being monitored prospectively at decided intervals. The study is also the first real world study generating safety data of COVAXIN in adolescents.

Nearly one third of selected adolescents developed AEFIs. Both local and systemic AEFIs were slightly more common after the second dose in adolescents. This is contrary to the trend observed by us as well as other groups in adults, where AEFIs decrease following the second dose.^11–13^ The observed incidence of local and systemic AEFIs in adolescents is higher than reported in the pre-print version of phase 2/3 trial of COVAXIN by Bharat Biotech (n=175).^4^ Close to 0.9% systemic AEFIs were of ‘severe’ grade compared to none in the phase 2/3 study. Both local and systemic reactogenicity rates observed in adolescents were higher than that reported previously in adults receiving COVAXIN.^13,14^ Apart from adolescents, a higher rate of occurrence of AEFIs (37-51%) was observed in adults too, contrary to the published studies showing close to 12-21% rates.^12,13^ Notwithstanding the higher rate of AEFIs, majority of these were mild-moderate and recovered over a median time of 1-2 days.

Regression analysis showed female adolescents and those with history of allergy to be at 1.5 times and 3 times higher risk of developing AEFIs. These two co-variates have recently been projected as risk factors of AEFI after COVAXIN in adults by a group from eastern India.^15^ Female sex, history of allergy and presence of hypothyroidism have been proposed as determinants of AEFI after COVISHIELD in adults.^11^ The association of AEFIs with hypothyroidism could not be examined in the present study because of a small number of individuals with thyroid disorders.

Other vaccines approved for adolescents include CoronaVac of Sinovac (China) and the BNT162b2 mRNA vaccine of Pfizer. Compared to what has been previously reported with CoronaVac, both local and systemic AEFIs were higher with COVAXIN in adolescents, particularly after the second dose.^16^

However, AEFI rates with BBV152 were lower compared to mRNA based vaccines with which high local (85%) and systemic (55-66%) reactogenicity rates have been observed.^17^ As in our study, systemic events have been reported to be more common after the second dose of mRNA vaccine. Among atypical adverse events, lymphadenopathy has been reported in 0.8% adolescent BNT162b2 recipients.^17^ In the current study, of the total interviewed adolescents six atypical AEFIs (0.9%) were reported. Among these, were two cases of increased bleeding which had recovered fully by the time of telephonic interview. Two atypical AEFIs were persisting till the time of interview and included a case of disturbing burning sensation in lower limbs and a case of aggravation of pre-vaccination skin allergy. The remaining two cases included recurrence of seizure after both doses in a female with pre-vaccination uncontrolled seizure disorder and hypothyroidism diagnosed within 3 weeks of the first dose in a female with pre-vaccination neck swelling. Both of these participants have been started on the necessary medications by their treating physicians and shall be monitored in the scheduled follow ups. In all, AEFIs were persistent in >1.5% adolescents, at the time of telephonic interview at day-14 and require further follow up to predict the course.

### 4.1. Limitations

Around half of the adolescent vaccinees were recruited at the time of receiving their second dose. Data of AEFIs following their first dose was collected at this time. The possibility of recall bias exists due to this study design. Telephonic follow-up at 14 days also may have introduced an element of recall bias. Since the study did not involve physical meetings and examinations of the enrolled participants, certain AEFIs such as vital sign variations may have been missed, leading to underestimation of AEFI incidence. Moreover, despite the definite questions asked on telephonic follow-up, individual vaccinee-specific factors may have resulted in minor variations in recording of AEFIs. The study also does not address the effectiveness aspect of the BBV152 vaccine in real world, the follow-up being short.

## 5. Conclusion

COVAXIN carries an overall favourable short-term safety profile in adolescents. The AEFI rates with COVAXIN are much lower compared to that reported with mRNA vaccines in adolescents. However, unlike in adults, AEFI rates were higher after the second dose which raises early questions on the effect of probable booster doses. Vigilance is needed while vaccinating females and those with a history of allergy. Watchfulness is also advised for bleeding events. A longer follow up as planned in the study may unravel more information about the long-term safety profile of COVAXIN in adolescents.

## Data Availability

Since this is a preliminary analysis, associated data may be made available by corresponding author on request.

## Funding

No funding support.

## Ethical Statement

No human experimentation performed. All procedures performed as per the Declaration of Helsinki and its further modifications. Written informed consent/assent obtained from all participants. Study conducted after permission from Institute Ethics Committee of the Institute of Medical Sciences, Banaras Hindu University.

## Declaration of competing interest

The authors have no conflicts of interest to declare.

## Acknowledgement

None.

## Notes

### Competing Interest Statement

The authors have declared no competing interest.

### Author Declarations

The study started after obtaining permission from the Ethics Committee of the Institute of Medical Sciences, Banaras Hindu University, Varanasi, UP, India.

