## Supplementary material for "A prospective observational study on BBV152 coronavirus vaccine use in adolescents and comparison with adults- first real-world safety analysis": Table 2

|  | Adolescents after first dose  (n=670) | Adolescents after second dose (n=340) | Adults after first dose  (n=181) | Adults after second dose (n=102) | Adults after booster dose  (n=12) |
| --- | --- | --- | --- | --- | --- |
| **Total AEFIs, n (%)** | **244 (36.3)** | **129 (37.9)** | **93 (51.4)** | **38 (37.2)** | **5 (41.7)** |
| **Local/ Systemic AEFIs^!^**  Only Local  Both Systemic and Local  Only Systemic | **142** (21.2)  **41** (6.1)  **61** (9.1) | **69** (20.3)  **27** (7.9)  **33** (9.7) | **53** (29.2)  **20** (11.0)  **20** (11.0) | **15** (14.7)  **4** (3.9)  **19** (18.6) | **0** (0)  **1** (8.3)  **4** (33.3) |
| **Severity of local AEFIs^!!^** |  |  |  |  |  |
| Mild | **152** (83.1) | **90** (93.7) | **71** (97.3) | **15** (78.9) | **1** (100.0) |
| Moderate | **31** (16.9) | **6** (6.3) | **2** (2.7) | **4** (21.1) | **0** (0) |
| **Severity of systemic AEFIs^!!!^** |  |  |  |  |  |
| Mild | **45** (44.1) | **27** (45.0) | **19** (47.5) | **13** (56.5) | **4** (80.0) |
| Moderate | **51** (50.0) | **33** (55.0) | **20** (50.0) | **10** (43.5) | **1** (20.0) |
| Severe | **6** (5.9) | **0** (0) | **1** (2.5) | **0** (0) | **0** (0) |
| **Individual AEFIs (LLT)^$^** |  |  |  |  |  |
| Injection site pain | **179** (26.7) | **96** (28.2) | **71** (39.2) | **18** (17.6) | **1** (8.3) |
| Fever | **82** (12.2) | **48** (14.1) | **29** (16.0) | **13** (12.7) | **1** (8.3) |
| Weakness | **13** (1.9) | **3** (0.9) | **7** (3.9) | **3** (2.9) | **1** (8.3) |
| Headache | **10** (1.5) | **11** (3.2) | **7** (3.9) | **4** (3.9) | **0** (0) |
| General body pain | **7** | **8** | **3** | **5** | **0** |
| Injection site swelling | **5** | **0** | **1** | **0** | **0** |
| Abdominal distress | **4** | **0** | **1** | **1** | **1** |
| Diarrhoea | **2** | **0** | **0** | **0** | **0** |
| Dizziness | **2** | **1** | **3** | **0** | **0** |
| Itching | **2** | **0** | **1** | **1** | **0** |
| Vomiting | **0** | **2** | **0** | **0** | **0** |
| Allergy (Skin allergy Increased) | **0** | **1** | **1** | **0** | **0** |
| Anorexia | **0** | **2** | **1** | **1** | **0** |
| Burning sensation in lower limbs | **1** | **0** | **0** | **0** | **0** |
| Cold (like features) | **1** | **0** | **1** | **0** | **0** |
| Convulsions* | **1** | **1** | **0** | **0** | **0** |
| Cough | **0** | **1** | **1** | **0** | **1** |
| Drowsiness | **0** | **1** | **0** | **0** | **0** |
| Dyspepsia (increased) | **0** | **0** | **1** | **0** | **0** |
| Dyspepsia | **0** | **0** | **1** | **0** | **0** |
| Epistaxis | **0** | **1** | **1** | **0** | **0** |
| Eye discomfort | **0** | **0** | **1** | **0** | **0** |
| Fall in blood pressure | **0** | **0** | **1** | **0** | **0** |
| Fatigue | **1** | **1** | **0** | **2** | **0** |
| Heavy menstrual bleeding | **1** | **0** | **1** | **0** | **0** |
| Hypothyroidism** | **1** | **0** | **0** | **0** | **0** |
| Joint pain (Upper limb) | **1** | **0** | **0** | **0** | **0** |
| Nausea | **1** | **1** | **1** | **0** | **0** |
| Pain of lower extremities | **1** | **0** | **0** | **0** | **0** |
| Throat sore | **0** | **0** | **0** | **0** | **1** |
| Unilateral leg swelling | **0** | **0** | **0** | **1** | **0** |
| **TTR (days)** | **0.5-7** | **0.5-14** | **1-12** | **1-7** | **2-7** |
| **AEFIs persisting at 14-days follow-up** | **3^&^** | **8** | **5** | **5** | **1** |

**Table 2: AEFIs in adolescents and adults after first, second and booster doses of BBV152 vaccine (COVAXIN)**

[*In a female with pre-vaccination uncontrolled seizure disorder, **In a female with pre-vaccination neck swelling

AEFI: adverse event following immunization, LLT: low level term, TTR: time to recovery

^!^All percentages are expressed with respect to total vaccinees in group

^!!^All percentages are expressed with respect to total number of local AEFIs

^!!!^All percentages are expressed with respect to total number of systemic AEFIs

^$^All percentages are expressed with respect to total vaccinees in group; percentages mentioned only for prominent AEFIs

^&^In one vaccinee, AEFI after first dose persisted till the time of second dose]

In
